# Comparative Effectiveness of Intravenous Ferric Carboxymaltose, Ferric Derisomaltose, and Iron Sucrose in Heart Failure With Reduced Ejection Fraction: A Network Meta-Analysis of 15 Randomized Trials

**DOI:** 10.1101/2025.09.11.25335472

**Authors:** Parag N. Patel, Maheen Adeeb, Sayyed Jalawan Asjad, Fnu Kalpana, Didar Singh, Devang Mangal

**Affiliations:** G.M.E.R.S. Medical College and Hospital, Patan, Gujarat, India; Jinnah Sindh Medical University, Karachi, Pakistan; SUNY Upstate Medical University, Syracuse, NY, USA; PT.B.D.Sharma Post Graduate Institute of Medical Sciences, Rohtak, Haryana, India; Springfield Memorial Hospital / Southern Illinois University, Springfield, USA; G.M.E.R.S Medical College and Hospital, Ahmedabad, Gujarat, India

## Abstract

**Background:** Iron deficiency affects nearly half of patients with heart failure with reduced ejection fraction and is associated with impaired functional capacity, recurrent hospitalizations, and increased mortality. Intravenous iron therapy improves exercise tolerance, but the comparative effectiveness of different formulations remains uncertain.

**Objectives:** To compare the efficacy of ferric carboxymaltose, ferric derisomaltose, and iron sucrose in patients with heart failure with reduced ejection fraction and iron deficiency using a network meta-analysis.

**Methods:** We systematically searched PubMed, the Cochrane Central Register of Controlled Trials, and Web of Science through August 2025 for randomized controlled trials of intravenous ferric carboxymaltose, ferric derisomaltose, or iron sucrose versus placebo. Fifteen randomized controlled trials published between 2007 and 2025 enrolling 7,761 patients were included. The primary outcomes were hospitalization for heart failure, all-cause mortality, and cardiovascular mortality. Secondary outcomes included change in six-minute walk distance, serum ferritin, and transferrin saturation. A frequentist random-effects network meta-analysis was performed, and treatment rankings were assessed using surface under the cumulative ranking curve probabilities.

**Results:** Twelve trials reported hospitalization for heart failure, showing significant reduction with ferric carboxymaltose (risk ratio 0.80, 95% confidence interval 0.69–0.92) and nonsignificant trends with ferric derisomaltose (0.80, 0.61–1.04) and iron sucrose (0.41, 0.15–1.07). No formulation reduced all-cause or cardiovascular mortality. Functional capacity improved with all formulations (mean difference +21.1 to +54.0 meters versus placebo), though heterogeneity was high. Ferritin and transferrin saturation increased significantly across all formulations, with the largest ferritin gain from ferric derisomaltose (+329 µg/L) and the most consistent transferrin saturation improvement from ferric carboxymaltose (+7.1%).

**Conclusions:** In patients with heart failure with reduced ejection fraction and iron deficiency, intravenous iron therapy—particularly ferric carboxymaltose—reduces hospitalization for heart failure, improves functional capacity, and corrects iron indices. Mortality benefits remain uncertain. These findings support guideline-endorsed use of intravenous iron, with ferric carboxymaltose as the best-studied option, while further outcome data for ferric derisomaltose and iron sucrose are needed.

## 1. Introduction

Iron deficiency (ID) affects ∼50% of patients with heart failure with reduced ejection fraction (HFrEF) and is an independent predictor of worse exercise capacity, higher hospitalization risk, and increased mortality [1]. Correcting ID has therefore become an important therapeutic target. Landmark trials such as FAIR-HF and CONFIRM-HF demonstrated that intravenous (IV) ferric carboxymaltose (FCM) improves symptoms, exercise tolerance, and quality of life in HFrEF [2], leading to its incorporation into international guidelines [3]. However, earlier studies were underpowered for clinical outcomes, leaving uncertainty about the effects of IV iron on hospitalizations and mortality.

Subsequent large trials produced mixed results. AFFIRM-AHF reported a 21% reduction in total HF hospitalizations and cardiovascular (CV) death [4], IRONMAN suggested fewer HF hospitalizations but was disrupted by the COVID-19 pandemic [5,7], and HEART-FID, the largest trial to date, did not meet its primary endpoint despite numerical reductions in HF hospitalizations and improved exercise capacity [6,9,10]. Recent systematic reviews and meta-analyses have clarified these findings: IV iron consistently reduces HF hospitalizations (∼20–25%) but has not shown a significant survival benefit [7,8,10,11]. Collectively, evidence supports IV iron as beneficial in reducing decompensation events and improving functional status, while mortality effects remain uncertain [7].

Most prior meta-analyses considered IV iron as a single class, despite differences among formulations. FCM is the most extensively studied and guideline-endorsed, ferric derisomaltose (FDI) was recently tested in the IRONMAN trial with results consistent in direction to FCM, and iron sucrose (IS) has been assessed only in small underpowered studies [5,12,13]. No head-to-head trials directly compare these formulations, though differences in dosing and pharmacology may influence clinical outcomes.

We therefore conducted a network meta-analysis of 15 RCTs in HFrEF with ID to compare three IV iron formulations—FCM, FDI, and IS—against placebo and each other. Our objectives were to assess their relative effects on HF hospitalizations, mortality, exercise capacity, and iron indices, and to establish a rank order of treatments to guide clinical decision-making.

## 2. Methods

### 2.1. Study design and protocol

We conducted a systematic review and network meta-analysis (NMA) in accordance with the PRISMA-NMA (Preferred Reporting Items for Systematic Reviews and Meta-Analyses – Network Meta-Analyses) guidelines. The study protocol was prospectively registered in PROSPERO (CRD4201142345). This was an investigator-initiated project with no external funding, and only aggregate trial-level data were analyzed.

### 2.2. Data sources and search strategy

We systematically searched PubMed/MEDLINE, the Cochrane Central Register of Controlled Trials (CENTRAL), and Web of Science from January 2000 to August 2025. The search strategy combined controlled vocabulary (MeSH) and free-text terms related to heart failure, iron deficiency, and intravenous iron supplementation.

No restrictions were applied for language or publication status; however, all included studies were available in English. The complete platform-specific search strategies are provided in Appendix 1.

### 2.3. Eligibility criteria and outcomes

Studies were eligible for inclusion if they: (i) were randomized controlled trials (RCTs) with a follow-up duration of at least 2 weeks; (ii) enrolled adult patients (≥18 years) with heart failure with reduced ejection fraction (HFrEF) and iron deficiency, defined as ferritin <100 µg/L or 100–300 µg/L with transferrin saturation <20%; (iii) compared intravenous (IV) iron therapy using ferric carboxymaltose (FCM), ferric derisomaltose (FDI), or iron sucrose (IS) with either placebo or standard care; and (iv) reported at least one of the prespecified outcomes of interest. We excluded trials of oral iron therapy, non-randomized studies, case reports, case series, editorials, reviews, and studies with insufficient outcome data.

Primary outcomes were heart failure hospitalization, all-cause mortality, and cardiovascular mortality, analyzed as risk ratios (RR) with 95% confidence intervals (CI). Secondary outcomes were change in 6-minute walk distance (6MWD), serum ferritin, and transferrin saturation (TSAT), expressed as mean differences (MD) with 95% CI. The definitions of all pooled outcomes are provided in Table S1.

### 2.4. Study selection and data extraction

Initially identified, 15 trials [4-6,12-23] all published between 2007 and 2025, satisfied inclusion after screening and full-text review (Figure 1).

**Figure 1.**
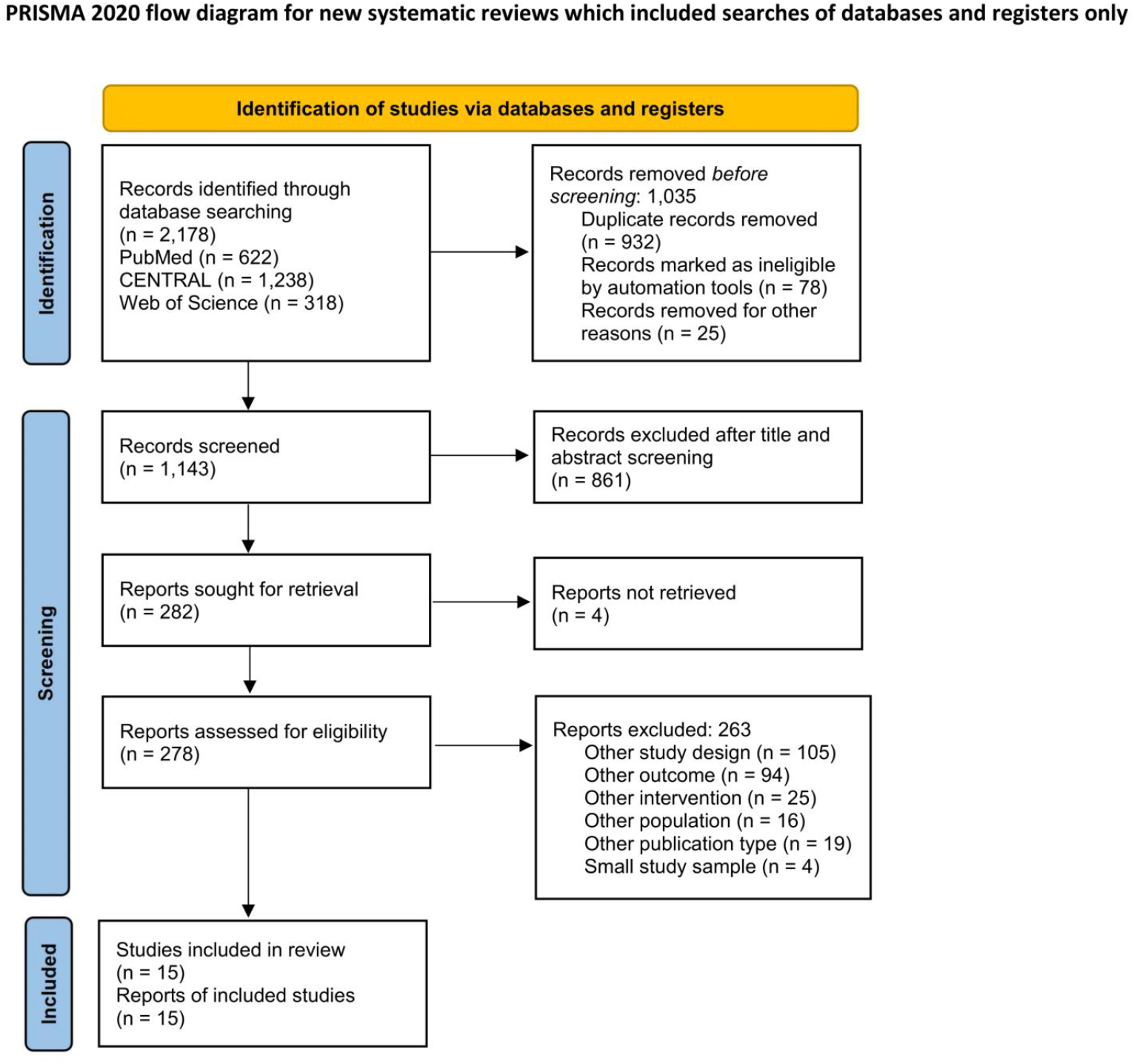
PRISMA flow diagram of study selection.

Two authors (P.P. and M.A.) independently extracted data from the included randomized controlled trials. Collected data included study characteristics (author, publication year, sample size), study population (patient demographics), description of interventions (drug name), and outcomes. Disagreements were resolved by discussion and consensus, with adjudication by a third author (F.K.) when necessary.

### 2.5. Quality assessment

Two authors (P.P. and S.A.) assessed the risk of bias using the Version 2 of the Cochrane Risk of Bias tool [24]. Risk of bias was assessed across five domains: randomization, deviations from intended variation, missing outcome data, measurement of outcome, and selection of reported results. The trials were scored as high, with some concerns, or low risk of bias in each domain.

### 2.6. Data analysis

We performed a frequentist pairwise meta-analysis using random-effects models, with heterogeneity quantified by the I^2^ statistic. A network meta-analysis was then conducted within a frequentist multivariate random-effects framework to integrate direct and indirect evidence across trials, accounting for multi-arm designs.

Treatment rankings were derived using surface under the cumulative ranking curve (SUCRA) probabilities, where higher values indicate greater likelihood of benefit. Consistency was evaluated qualitatively, as the network was star-shaped with placebo as the common comparator, and sensitivity analyses explored the influence of individual large trials. Certainty of evidence for each outcome was assessed using the GRADE framework adapted for network meta-analysis. In global inconsistency testing, p < 0.05 was considered to indicate statistically significant heterogeneity. For the node-splitting approach, p < 0.05 indicated a statistically significant inconsistency between direct and indirect evidence.

All analyses were performed in R (netmeta package), and results are presented as network plots, pooled forest plots, league tables, and SUCRA rankings.

## 3. Results

### 3.1. Study selection

The database search yielded 2,178 records, of which 1,035 were removed as duplicates or ineligible before screening. After title/abstract screening and full-text review, 15 randomized controlled trials (RCTs) were included (Figure 1). These studies enrolled patients with HFrEF and iron deficiency and compared intravenous ferric carboxymaltose (FCM), ferric derisomaltose (FDI), or iron sucrose (IS) with placebo or standard care.

### 3.2. Study characteristics

Fifteen RCTs [4–6,12–23] were pooled in our analysis, with a total of 7,761 participants. The publication years of the included trials ranged from 2007 to 2025. The number of patients in the treatment groups who received intravenous (IV) iron was 3,971, and the number of patients in the control groups was 3,790. The studies varied in sample sizes, type of IV iron administered, baseline LVEF%, and follow-up durations. The participants’ mean age ranged from 51 to 75.4 years, with males making up the majority of the study population in 13 studies. The mean follow-up duration was 33.96 weeks. Nine studies with 6,433 patients used ferric carboxymaltose [4,6,14,17-19,21-23], three studies with 151 patients used iron sucrose [12,13,15,16], and two studies with 1,177 patients used ferric derisomaltose [5,20]. The detailed characteristics of the included studies and patients are presented in Table 1, while baseline clinical and demographic information is shown in Table 2.

**Table 1.** Characteristics of included randomized controlled trials.

|  | Country | Sample Size (Total) | Definition of Iron Deficiency | Inclusion Criteria | Follow-up Duration | Primary Endpoint | Secondary Endpoint |
| --- | --- | --- | --- | --- | --- | --- | --- |
| Toblli et al., 2007 [12] | Argentina | 40 | Ferritin<100 ng/mL, TSAT< 20%, Hb <12.5 g/dl for men and <11.5 g/dl for women | NYHA II to IV; CrCl 90 mL/min | 24 weeks | Change in NT-proBNP level and CRP, Hb increase, TSAT and Ferritin increase, renal parameter bAQ:9; creatinine clearance (change in hematologic and renal parameter) | NYHA, MLHFQ, 6MWT, Hospitalizations |
| FERRIC-HF (Okonko et al., 2008) [13] | UK, Poland, Germany | 35 | Ferritin <100 µg/L or 100–300 µg/L with TSAT <20% | NYHA II or III (LVEF ≤45%) ; Peak VO<sub>2</sub> ≤18 mL/kg/min ; Hb ≤12.5 g/dL (anemic) or 12.5–14.5 g/dL (non-anemic) | 18 weeks | Change in absolute peak VO<sub>2</sub> (mL/min) | NYHA class, exercise duration, MLHFQ, fatigue score, TSAT, ferritin, Hb, LVEF, safety |
| FAIR-HF (Anker et al., 2009) [14] | 11 countries (Russia, Ukraine, Poland) | 459 | Ferritin <100 µg/L, or 100–299 µg/L with TSAT <20% | LVEF ≤40% (NYHA II) or ≤45% (NYHA III) ; Hb 95–135 g/L | 24 weeks | Patient Global Assessment (PGA) and NYHA class at Week 24 | 6MWT distance, KCCQ score, EQ-5D VAS |
| IRON-HF (Beck-da-Silva et al., 2013) [15] | Brazil | 23 | Ferritin <500 ng/mL with TSAT <20% | Stable outpatients with HF for ≥3 months; NYHA II–IV ; LVEF <40% ; Hb 9–12 g/dL | 12 weeks | Change in peak VO<sub>2</sub> (mL/kg/min) | Hb increase, TSAT >20%, anemia correction, ferritin change |
| Toblli et al., 2015 [16] | Argentina | 60 | Ferritin <100 ng/mL, or 100–300 ng/mL with TSAT <20%; Hb <12.5 g/dL (men), Hb <11.5 g/dl (women) | LVEF 35%, NYHA class II–IV, CrCl 90 mL/min | 24 weeks | LVEF in two- and four-chamber | Change in Hb, ferritin and TSAT, CRP and NT-pro-BNP |
| CONFIRM-HF (Ponikowski et al., 2015) [17] | 9 countries (Russia, Ukraine, Poland) | 301 | Ferritin <100 ng/mL, or 100–300 ng/mL with TSAT <20% | - NYHA II or III (LVEF ≤45%) ; BNP >100 pg/mL or NT-proBNP >400 pg/mL ; Hb <15 g/dL | 52 weeks | Change in 6-minute walk test (6MWT) at 24 weeks | NYHA class, PGA, fatigue score, QoL scores (KCCQ, EQ-5D), HF hospitalizations, death |
| EFFECT-HF (van Veldhuisen et al., 2017) [18] | 9 countries (NR) | 174 | Ferritin <100 ng/mL, or 100–300 ng/mL with TSAT <20% | NYHA II–III (LVEF ≤45%) ; Peak VO<sub>2</sub> 10–20 mL/kg/min ; BNP >100 pg/mL or NT-proBNP >400 pg/mL | 24 weeks | Change in peak VO<sub>2</sub> (mL/kg/min) | TSAT, ferritin, Hb, BNP/NT-proBNP, NYHA class, PGA, VE/VCO<sub>2</sub> slope, hospitalizations, safety |
| PRACTICE-ASIA-HF (Yeo et al., 2018) [19] | Singapore | 50 | Ferritin <300 ng/mL or TSAT <20% | Hospitalized patients for acute decompensated HF, ferritin <300 ng/mL, TSAT% <20% | 12 weeks | 6MWT at 12 weeks | Quality of life: KCCQ, Visual Analogue Scale (VAS) |
| FERRIC-HF II (Edwards et al., 2019) [20] | London | 40 | Ferritin <100 µg/L or 100–300 µg/L with TSAT <20% | NYHA II or III; LVEF ≤45%, Use of optimal HF drugs for ≥4 weeks without dose changes for ≥2 weeks | 2 weeks | Skeletal muscle energetics PCr t1/2 at 2 weeks | ADP recovery half-time (ADP t1/2; energetic marker), 6MWT, Peak VO<sub>2</sub>, Ratio of Minute Ventilation to CO<sub>2</sub>, symptoms quantified by NYHA Class, Borg dysnea score, fatigue scale. KCCQ, HB, Iron status (ferritin, TSAT), N-terminal pro-B-type natriuretic peptide; and LVEF. |
| AFFIRM-AHF (Ponikowski et al., 2020) [4] | 15 countries (Georgia, Ukraine, Romania) | 1108 | Ferritin <100 ng/mL, or 100–299 ng/mL with TSAT <20% | Hospitalized for acute HF ; LVEF <50% ; Received ≥40 mg IV furosemide ; NT-proBNP ≥1600 pg/mL (SR) or ≥2400 pg/mL (AF) ; Hb 80-150 g/L | 52 weeks | Composite of total HF hospitalizations and cardiovascular dea | Total CV hospitalizations + CV death, CV death alone, time to first HF hospitalization or CV death, days lost due to HF hospitalizations or CV death |
| FCM-HF-IN (Dhoot et al., 2020) [21] | India | 70 | Hb <12 g/dl women and <13 g/dl men. | NYHA Functional Class II/III | 12 weeks | Change in peak VO<sub>2</sub> (mL/kg/min) | Change in levels of ferritin, hemoglobin (Hb), left ventricular ejection fraction, 6MWT, NYHA Class and the Minnesota Living with Heart Failure Questionnaire (MLHFQ) score |
| IRON-CRT (Martens et al., 2021) [22] | Belgium | 75 | Ferritin <100 ng/mL, or 100–299 ng/mL with TSAT <20% | LVEF <45%; NYHA Functional Class II/III, > 98% biventricular pacing the last 6 months | 12 weeks | Change in LVEF | Change from baseline in left ventricular end- systolic (LVESV) and end-diastolic volume (LVEDV); cardiopulmonary exercise test variables of peak oxygen consumption (VO<sub>2</sub>) and the slope of minute ventilation/carbon dioxide production (VE/VCO<sub>2</sub>) ratio, Kansas City Cardiomyopathy Questionnaire (KCCQ) score; N-terminal pro B-type natriuretic peptide (NT-proBNP) level. |
| IRONMAN (Kalra et al., 2022) [5] | UK | 1137 | Ferritin <100 µg/L or TSAT <20% | LVEF ≤45% within prior 24 months ; Current or recent (<6 m) HF hospitalisation | Median ~2.7 years (IQR 1.8–3.6) | Composite of recurrent HF hospitalizations and cardiovascular death | CV hospitalization (first event), CV death, MLHFQ and EQ-5D scores, 6MWT (4 & 20 mo), all-cause mortality, HF rehospitalization, safety endpoints (infection, blood transfusion, etc.) |
| HEART-FID (Mentz et al., 2023) [6] | 14 countries (North America, EU, Russia/Ukraine) | 3065 | Ferritin <100 ng/mL, or 100–300 ng/mL with TSAT <20%; Hb >9 g/dL and <13.5 (F) or <15 (M) | HF with LVEF ≤40% | Median 1.9 (1.3-3.0) | Death at 12 months, HF hospitalization at 12 months<br>Change in 6MWD at 6 month | CV death or HF hospitalization , MLHFQ, EQ-5D |
| FAIR-HF2 (Anker et al., 2025) [23] | UK | 1105 | Ferritin <100 ng/mL, or 100–299 ng/mL with TSAT <20% | HF with LVEF ≤45% | Median 16.6 months (IQR,7.9-29.9 months) | Time to first event cardiovascular death or heart failure hospitalization; recurrent heart failure hospitalizations and time to first event of either cardiovascular death or heart failure hospitalization in sub- group of TAST 20% at baseline | Change NYHA class, EQ-5D score ,and 6-minute walk distance, global assessment of subjective well-being score |

**Table 2.** Baseline demographics of enrolled patients.

| Study | Arm | Sample Size (per arm) | Iron Form / Class | Age (Mean ± SD) | NYHA Class Distribution (I/II/III/IV) | Hemoglobin (g/dL) | Ferritin (ng/mL) | TSAT (%) | BNP or NT-proBNP Criteria |
| --- | --- | --- | --- | --- | --- | --- | --- | --- | --- |
| Toblli et al., 2007 [12] | IV Iron | 20 | IV Iron Sucrose | 76 ± 7 years | NYHA functional class: 2.9 ± 0.7 | 10.3 ± 0.6 | 73.0 ± 29.9 | 0.20 ± 0.01 | 255.9 ± 124.6 (NT) |
|  | Placebo | 20 |  | 74 ± 8 years | NYHA functional class: 2.9 ± 0.6 | 10.2 ± 0.5 | 70.6 ± 21.4 | 0.20 ± 0.01 | 267.5 ± 114.9 |
| FERRIC-HF (Okonko et al., 2008) [13] | IV Iron | 24 | IV Iron Sucrose | 64 ± 14 years | 0/13/11/0 | ≤12.5 g/dL | <100 or 100–300 | <20% | NR |
|  | Placebo | 11 |  | 62 ± 11 years | 0/6/5/0 | 12.5–14.5 | <100 or 100–300 <20% | <20% | NR |
| FAIR-HF (Anker et al., 2009) [14] | IV Iron | 304 | IV FCM | 67.8 ± 10.3 years | 0/53/251/0 | 11.9 ± 1.3 | 52.5 ± 54.5 | 17.7 ± 12.6 | NR |
|  | Placebo | 155 |  | 67.4 ± 11.1 years | 0/29/126/0 | 11.9 ± 1.4 | 60.1 ± 66.5 | 16.7 ± 8.4 | NR |
| IRON-HF (Beck-da-Silva et al., 2013) [15] | IV Iron | 10 | IV Iron Sucrose | 66.9 ± 8.3 years | Not reported | 11.2 ± 0.6 | 185 ± 146 | 18.9 ± 9.7 | NR |
|  | Placebo | 6 |  | 68.9 ± 10.1 years | Not reported | 10.9 ± 0.7 | 95 ± 128 | 13.5 ± 5.8 | NR |
| Toblli et al., 2015 [16] | IV Iron | 30 | IV Iron Sucrose | 75.4 ± 6 years | 3.0 ± 0.7 | 10.1 ± 0.8 | 70.6 ± 24.9 | 19.2 ± 1.8 | 366.0 ± 200.0 (NT pro) |
|  | Placebo | 30 |  | 74.7 ± 7 years | 3.1 ± 0.6 | 10.1 ± 0.6 | 68.4 ± 18.3 | 19.2 ± 1.9 | 378.0 ± 195.1 |
| CONFIRM-HF (Ponikowski et al., 2015) [17] | IV Iron | 150 | IV FCM | 68.8 ± 9.5 years | 0/80/70/0 | 12.37 ± 1.41 | 57.0 ± 48.4 | 20.2 ± 17.6 | BNP >100 pg/mL or NT-proBNP >400 pg/mL |
|  | Placebo | 151 |  | 69.5 ± 9.3 years | 0/91/60/0 | 12.42 ± 1.30 | 57.1 ± 41.6 | 18.2 ± 16.1 | BNP >100 pg/mL or NT-proBNP >400 pg/mL |
| EFFECT-HF (van Veldhuisen et al., 2017) [18] | IV Iron | 88 | IV FCM | 63 ± 12 years | 0/61/25/0 | 12.9 ± 1.3 | 48 (median) | 17.3 (median) | BNP >100 pg/mL or NT-proBNP >400 pg/mL |
|  | Placebo | 86 |  | 64 ± 11 years | 0/54/32/0 | 13.0 ± 1.5 | 53 (median) | 18.1 (median) | BNP >100 pg/mL or NT-proBNP >400 pg/mL |
| PRACTICE-ASIA-HF (Yeo et al., 2018) [19] | IV Iron | 24 | IV FCM | 61.1 ± 10.8 years | Not reported | 11.6 ± 1.9 | 91.4 (80.4) | 15.7 (10.1) | NR |
|  | Placebo | 25 |  | 64 ± 10 years | Not reported | 13.1 ± 1.3 | 84.1 (63.7) | 13.9 (6.8) | NR |
| FERRIC-HF II (Edwards et al., 2019) [20] | IV Iron | 21 | IV Ferric Derisomaltose | 70 ± 12 years | 0/12/9/0 | 130 ± 15 | 34 (18–50) | 21 ± 8 | NT BNP 1486 (245–2054) |
|  | Placebo | 19 |  | 62 ± 13 years | 0/9/10/0 | 128 ± 20 | 59 (39–79) | 18 ± 10 | NT BNP 462 (206–855) |
| AFFIRM-AHF (Ponikowski et al., 2020) [4] | IV Iron | 567 | IV FCM | 71.2 ± 10.8 years | 14/255/277/6 | 12.3 ± 1.6 | 83.9 ± 62.2 | 15.2 ± 8.3 | BNP median: 1068 pg/mL NT-proBNP median: 4743 pg/mL (IQR 2781–8128) |
|  | Placebo | 565 |  | 70.9 ± 11.1 years | 8/240/276/22 | 12.1 ± 1.6 | 88.5 ± 68.8 | 14.2 ± 7.5 | BNP median: 1204 pg/mL NT-proBNP median: 4684 pg/mL (IQR 2785–8695) |
| FCM-HF-IN (Dhoot et al., 2020) [21] | IV Iron | 35 | IV FCM | 51.0 ± 11.6 years | 0/26/9/0 | 11 ± 1.4 | 40.1 ± 27.2 | NR | NR |
|  | Placebo | 35 |  | 54.8 ± 9.0 years | 0/30/5/0 | 11.3 ± 0.9 | 45.5 ± 35.1 | NR | NR |
| IRON-CRT (Martens et al., 2021) [22] | IV Iron | 37 | IV FCM | 72 ± 12 years | 0/22/15/0 | 13.3 ± 1.2 | 82 [38–106] (median) | 18.8 ± 6.0 | NT-proBNP median 2227 [299–2967] |
|  | Placebo | 38 |  | 73 ± 9 years | 0/19/19/0 | 13.1 ± 1.3 | 81 [43–99] (median) | 19.4 ± 7.0 | NT-proBNP median 1604 [767–2204] |
| IRONMAN (Kalra et al., 2022) [5] | IV Iron | 569 | IV Ferric Derisomaltose | 73.2 (66.7–80.1) (Median [IQR]) | 0/328/230/11 | 12.1 (IQR 11.2–12.8) (Median) | 49.0 (30.0–86.0) (median) | 15% (11–20) (median) | NT-proBNP >250 (SR) or >1000 (AF), BNP >75 (SR) or >300 (AF) |
|  | Placebo | 568 |  | 73.5 (67.1–79.1) (Median [IQR]) | 0/320/238/10 | 12.1 (IQR 11.2–12.9) (Median) | 50.0 (30.0–85.0) (median) | 15% (10–19) (median) | NT-proBNP >250 (SR) or >1000 (AF), BNP >75 (SR) or >300 (AF) |
| HEART-FID (Mentz et al., 2023) [6] | IV Iron | 1532 | IV FCM | 68.6 ± 10.9 years | 0/797/711/22 | 12.6 ± 1.4 | 56.0 ± 47.3 | 23.9 ± 11.2 | NT-proBNP median: 1485.5 (IQR: 727.1–3044.5) |
|  | Placebo | 1533 |  | 68.6 ± 11.2 years | 0/820/692/19 | 12.5 ± 1.4 | 12.5 ± 1.4 | 23.0 ± 10.3 | NT-proBNP median: 1423.6 (IQR: 710.0–2883.8) |
| FAIR- HF2 (Anker et al., 2025) [23] | IV Iron | 558 | IV FCM | 70.1 ± 11.4 | 0/369/186/1 | 0/359/184/3 | 72 ± 52 | 18.6 ± 9.3 | NT-proBNP median 1734 (834-5007) |
|  | Placebo | 547 |  | 69.7 ± 12.0 |  |  | 74 ± 58 | 17.9 ± 9.0 | NT-proBNP median 2022 (841-4825) |

### 3.3. Network of evidence

The treatment network for all outcomes was star-shaped, with placebo serving as the common comparator (Figure 2). No head-to-head comparisons between active intravenous iron formulations were available; therefore, indirect comparisons were derived through the placebo anchor.

**Figure 2.**
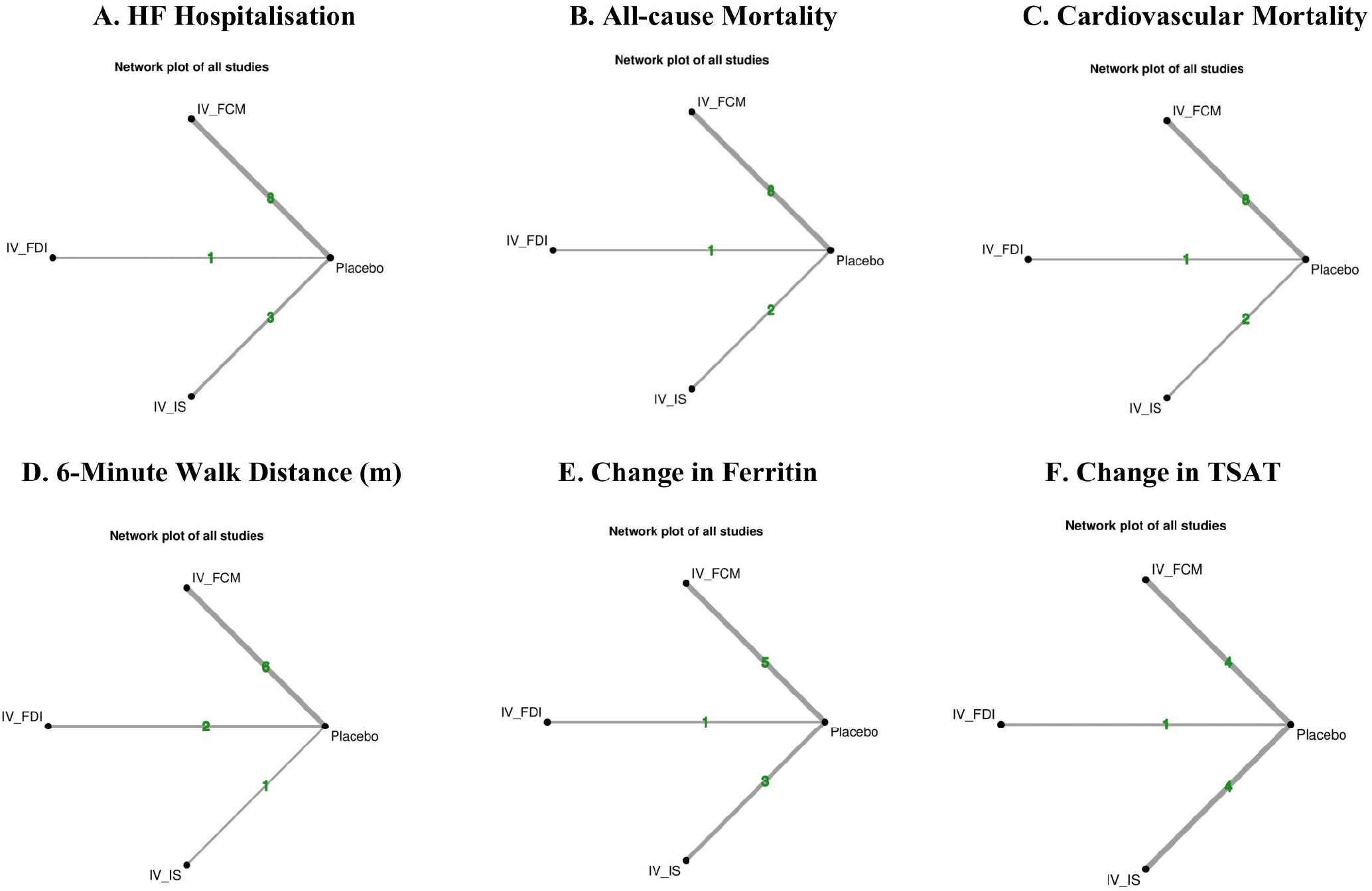
Network geometry of included interventions across outcomes.

### 3.4. Risk of bias

Twelve studies had an overall low risk of bias. The remaining three studies had some concerns of bias. The details of the bias assessment for each included study are shown in Figure S1.

### 3.5. Heart failure hospitalization

Twelve RCTs (IV FCM 8, IV FDI 1, IV IS 3) including a total of 7,632 patients reported data on HF hospitalization. In the pooled network meta-analysis, intravenous ferric carboxymaltose (IV FCM) significantly reduced the risk of HF hospitalization compared with placebo (RR 0.80, 95% CI 0.69–0.92). Ferric derisomaltose (IV FDI: RR 0.80, 95% CI 0.61–1.04) and iron sucrose (IV IS: RR 0.41, 95% CI 0.15–1.07) showed nonsignificant trends in the same direction (Figure 3A). Between-study heterogeneity was moderate (I^2^ = 49.1%), with tau^2^ estimated at 0.0002, indicating low absolute variance in treatment effects. The treatment hierarchy favored IV IS (SUCRA 93%), followed by FCM (51%), FDI (47%), and placebo (7%) (Figure 4A). League tables confirmed the superiority of active formulations over placebo (Figure 5A). The summary forest plot (Figure 6A) consistently ranked IV IS and FCM as the most effective interventions for reducing HF hospitalization. Publication bias was explored with funnel plots, which showed no major asymmetry (Figure S2). Meta-regression analyses did not identify significant effect modification across trials (Figure S4).

**Figure 3.**
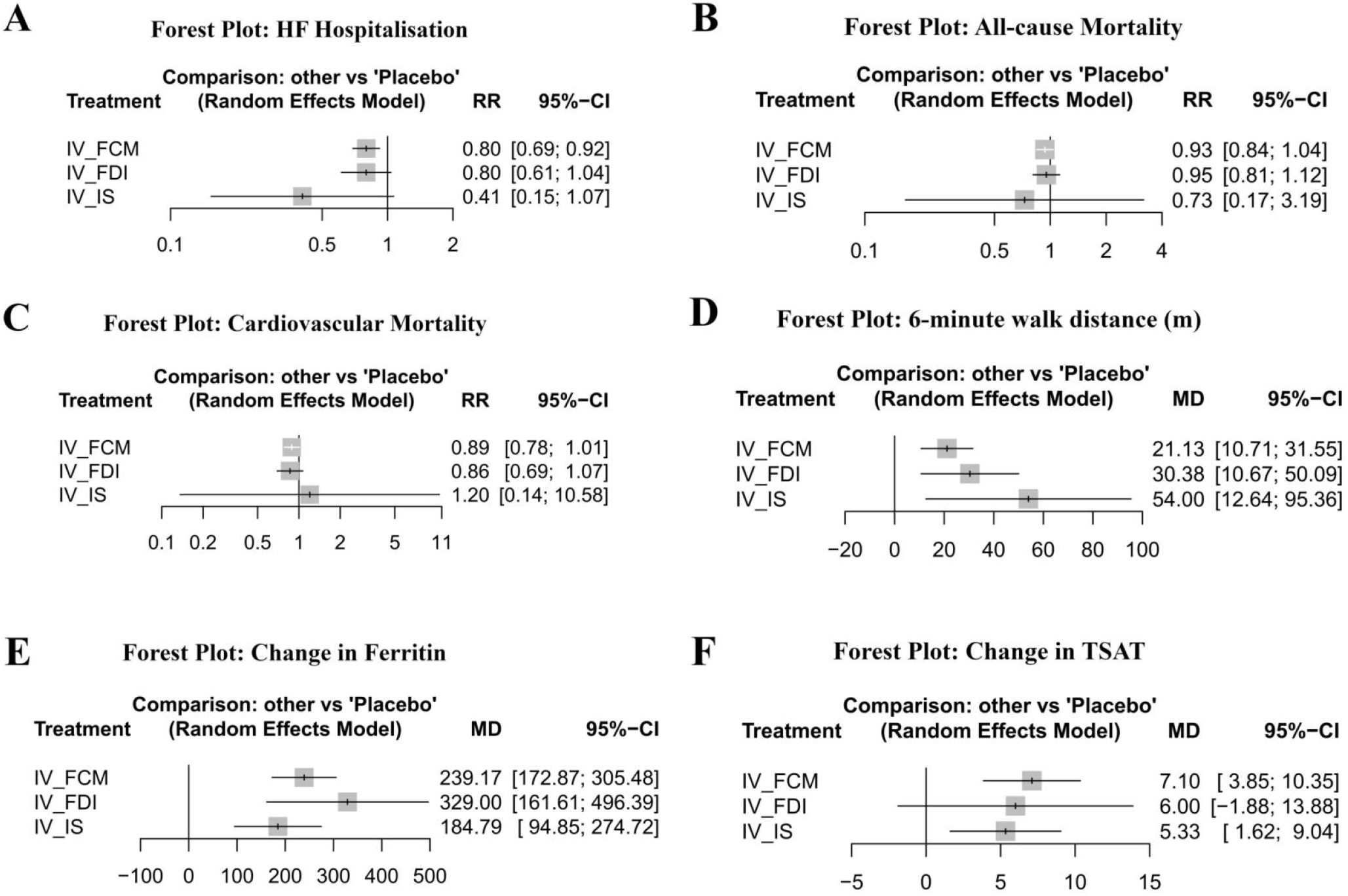
Forest plots of pooled network meta-analysis estimates:

**Figure 4.**
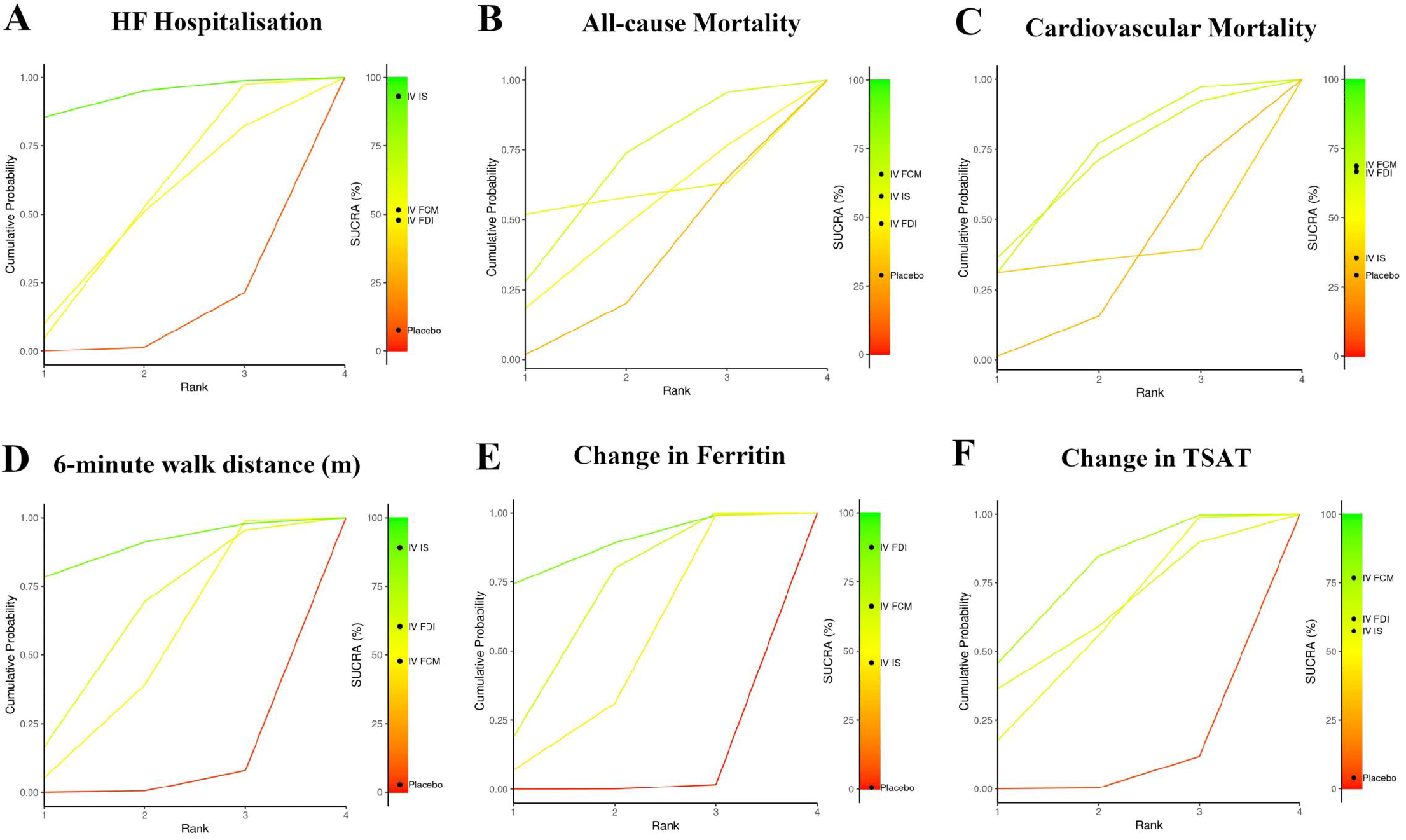
SUCRA ranking plots for all outcomes (A–F).

**Figure 5.**
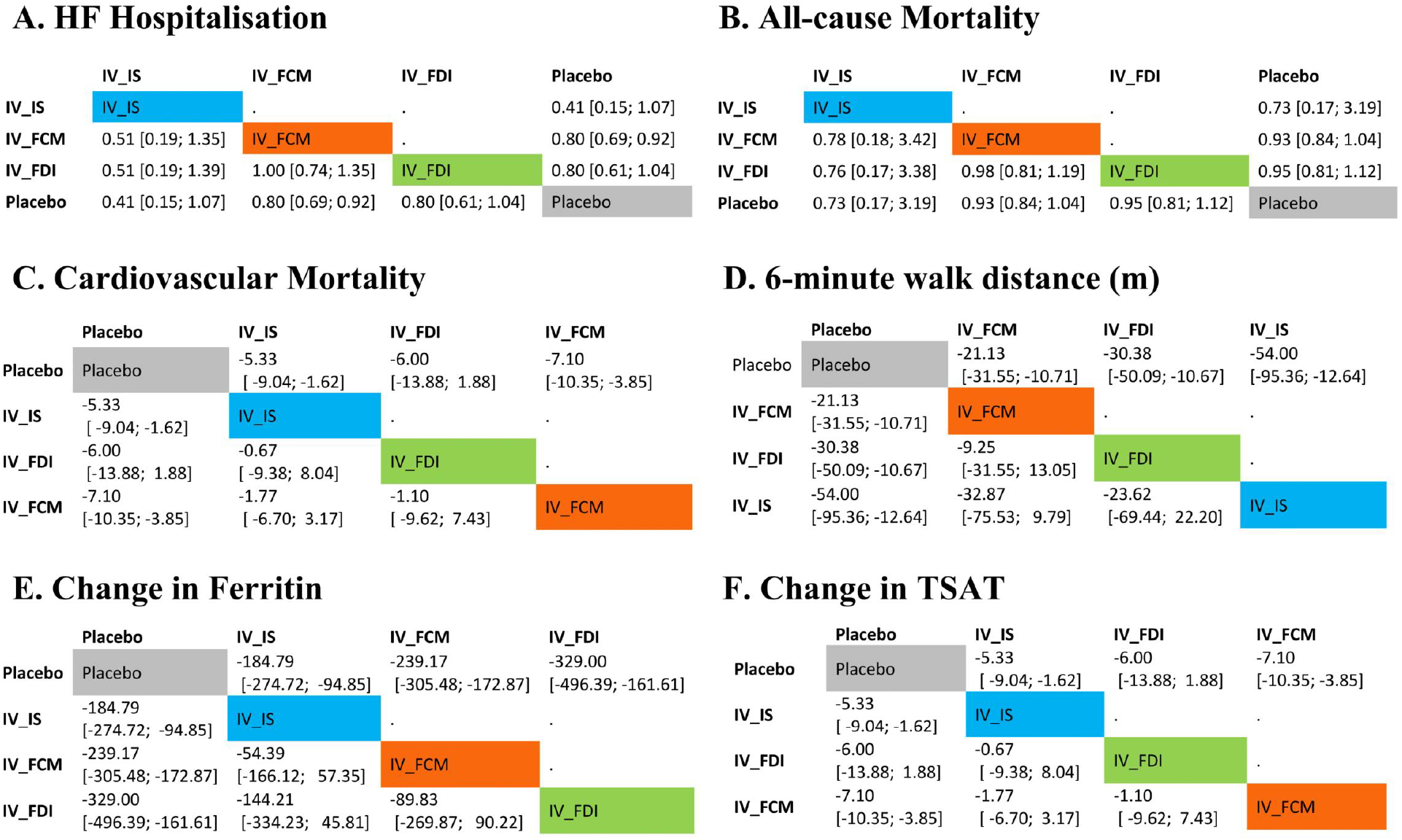
League tables of all pairwise comparisons (A–F).

**Figure 6.**
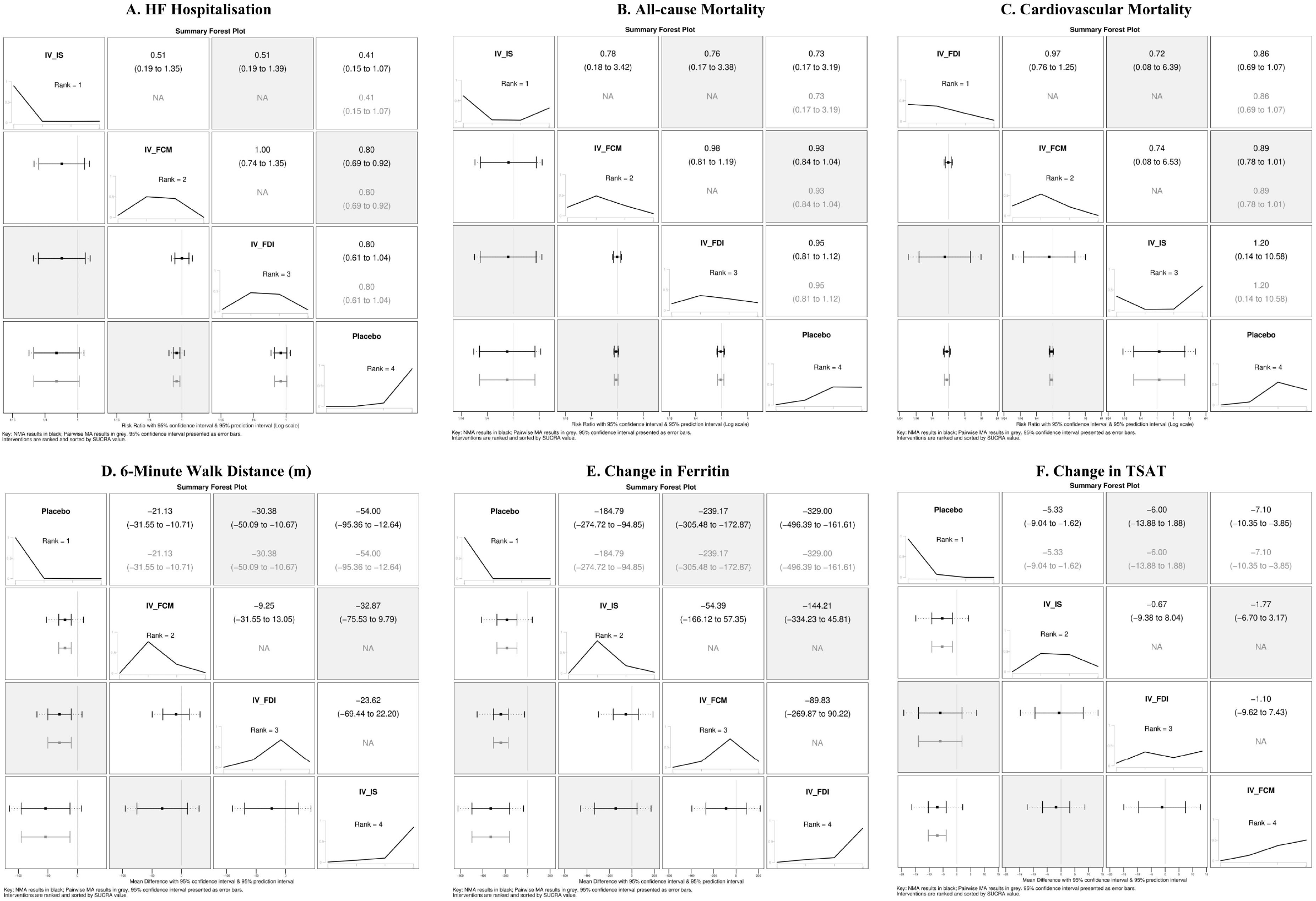
Summary forest plots across outcomes (A–F).

### 3.6. All-cause mortality

Eleven RCTs (IV FCM 8, IV FDI 1, IV IS 2) with 7,618 patients reported all-cause mortality outcomes. In the pooled network meta-analysis, no intravenous iron formulation significantly reduced mortality compared with placebo (Figure 3B). Risk ratios were 0.93 (95% CI 0.84–1.04) for ferric carboxymaltose (FCM), 0.95 (95% CI 0.81–1.12) for ferric derisomaltose (FDI), and 0.73 (95% CI 0.17–3.19) for iron sucrose (IS). Between-study heterogeneity was negligible (I^2^ = 0%, tau^2^ <0.0001). Ranking probabilities suggested modest advantages for active treatments compared with placebo, though differences were not clinically meaningful. SUCRA values were 65.8% for FCM, 57.7% for IS, 47.6% for FDI, and 28.9% for placebo (Figure 4B). League tables (Figure 5B) showed overlapping confidence intervals across all active treatments, indicating no clear superiority of one formulation over another. The summary forest plot (Figure 6B) confirmed the overall neutral effect on mortality. Funnel plots did not reveal major asymmetry, and meta-regression analyses found no significant effect modification (Figure S2, S4).

### 3.7. Cardiovascular mortality

Eleven RCTs (IV FCM 8, IV FDI 1, IV IS 2) with a total of 7,543 patients reported cardiovascular mortality. In the pooled NMA, none of the intravenous iron formulations significantly reduced cardiovascular death compared with placebo (Figure 3C). Risk ratios were 0.89 (95% CI 0.78–1.01) for ferric carboxymaltose (FCM), 0.86 (95% CI 0.69–1.07) for ferric derisomaltose (FDI), and 1.20 (95% CI 0.14–10.58) for iron sucrose (IS). Between-study heterogeneity was negligible (I^2^ = 0%, tau^2^ <0.0001). Treatment rankings suggested modest advantages of FCM and FDI, though not statistically meaningful. SUCRA values were 68.6% for FCM, 66.7% for FDI, 35.4% for IS, and 29.3% for placebo (Figure 4C). League tables confirmed overlapping estimates across all active treatments (Figure 5C), and the summary forest plot (Figure 6C) reinforced the overall neutral effect. Funnel plot inspection did not indicate significant publication bias, and meta-regression analyses showed no evidence of effect modification (Figure S2, S4).

### 3.8. 6-Minute Walk Distance (6MWD)

Nine randomized trials (IV FCM 6, IV FDI 2, IV IS 1) including 6,266 patients evaluated change in 6MWD (Figure 3D). All three intravenous iron formulations significantly improved functional capacity compared with placebo. The mean differences versus placebo were +21.1 m (95% CI 10.7–31.6) for ferric carboxymaltose (FCM), +30.4 m (95% CI 10.7–50.1) for ferric derisomaltose (FDI), and +54.0 m (95% CI 12.6–95.4) for iron sucrose (IS). Between-study heterogeneity was substantial (τ^2^ = 188.9; I^2^ = 96.8%), reflecting variability across trial designs and patient populations. Ranking analysis suggested that iron sucrose had the highest probability of being most effective (SUCRA 89.0%), followed by FDI (60.4%) and FCM (47.7%), whereas placebo consistently ranked worst (SUCRA 2.8%) (Figure 4D). League tables confirmed overlapping estimates across all active treatments (Figure 5D). Funnel plots suggested potential small-study effects, but overall results were directionally consistent (Figure S2).

### 3.9 Change in Iron Indices

Nine RCTs (IV FCM 5, IV FDI 1, IV IS 3) enrolling 4,923 patients reported changes in serum ferritin. Compared with placebo, all intravenous formulations significantly increased ferritin: IV ferric carboxymaltose (MD 239.2 µg/L, 95% CI 172.9–305.5), IV ferric derisomaltose (MD 329.0 µg/L, 95% CI 161.6–496.4), and IV iron sucrose (MD 184.8 µg/L, 95% CI 94.9–274.7) (Figure 3E). While between-treatment comparisons were not statistically significant, IV ferric derisomaltose yielded the largest point estimate. SUCRA rankings placed IV ferric derisomaltose as most effective (87.5%), followed by IV ferric carboxymaltose (66.2%), IV iron sucrose (45.7%), and placebo (0.6%) (Figure 4E). Heterogeneity across trials was high (τ^2^ = 7149.3; I^2^ = 99.1%), reflecting variability in trial populations and follow-up periods. Funnel plots suggested possible small-study effects, though meta-regression analyses did not reveal consistent effect modification (Figure S2, S4).

Similarly, nine RCTs (IV FCM 4, IV FDI 1, IV IS 4) with 4,994 patients evaluated transferrin saturation (TSAT). All three intravenous formulations improved TSAT compared with placebo: IV ferric carboxymaltose (MD 7.10%, 95% CI 3.85–10.35), IV ferric derisomaltose (MD 6.00%, 95% CI −1.88 to 13.88), and IV iron sucrose (MD 5.33%, 95% CI 1.62–9.04) (Figure 3F). Ranking analysis indicated IV ferric carboxymaltose had the highest probability of benefit (SUCRA 76.7%), followed by IV ferric derisomaltose (61.8%), IV iron sucrose (57.4%), and placebo (4.0%) (Figure 4F). Considerable heterogeneity was present (τ^2^ = 9.78; I^2^ = 94.8%), though funnel plot inspection did not reveal strong evidence of small-study effects (Figure S2).

A GRADE Summary of Findings table (Table 3) provides an overview of the certainty of evidence for all outcomes assessed in this network meta-analysis. The approach for constructing and interpreting the Summary of Findings table was based on established GRADE guidance for network meta-analyses and methodological resources provided by Cochrane [25,26].

**Table 3.**
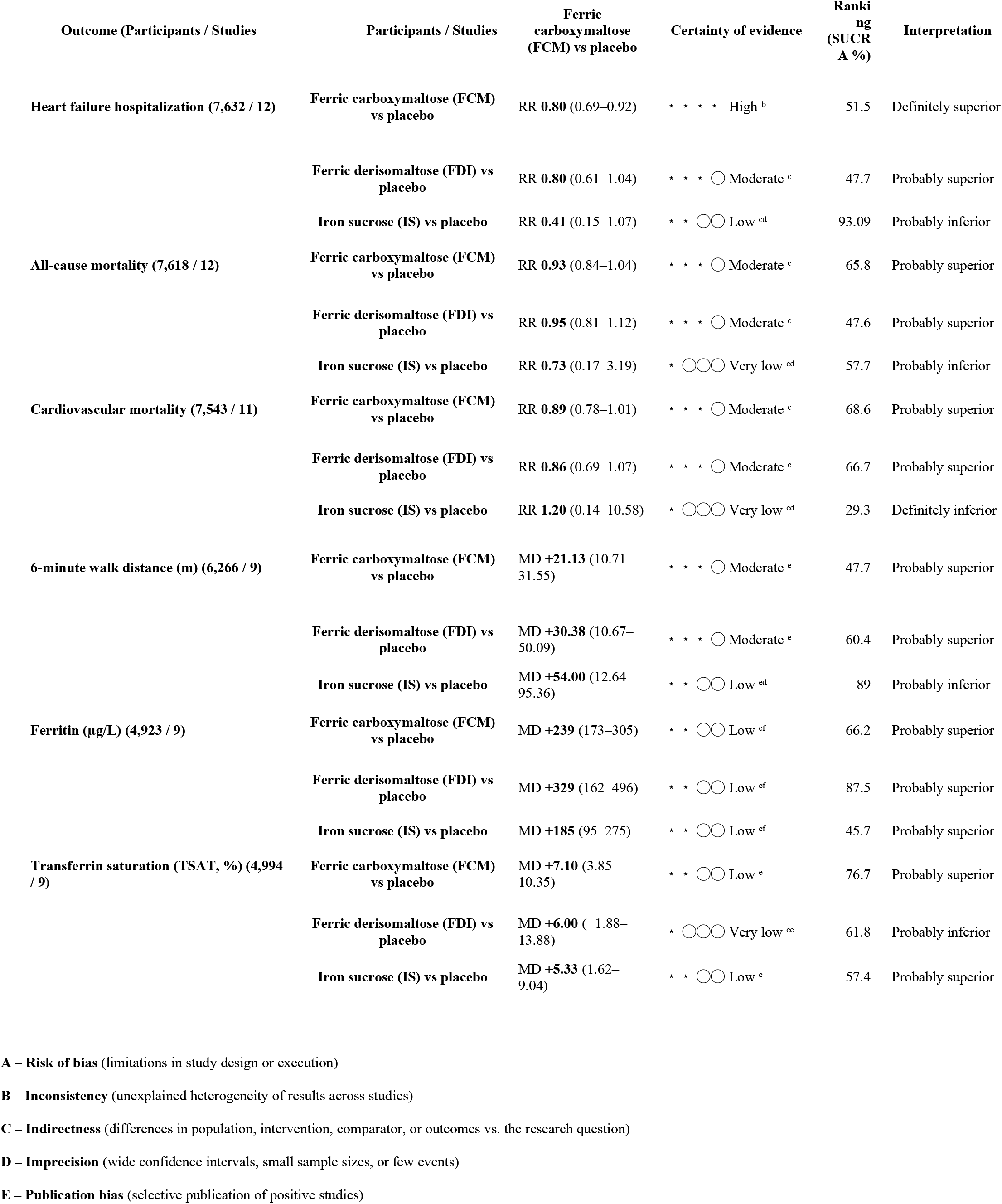
Summary of findings with GRADE assessment.

## 4. Discussion

This network meta-analysis synthesized evidence from 15 randomized controlled trials including more than 7,761 patients with HFrEF and iron deficiency. We specifically compared FCM, FDI, and IS because they are the three most commonly used intravenous iron formulations in HFrEF. These agents differ in pharmacologic structure, dosing regimens, and availability across regions. FCM allows large single doses and has the largest RCT evidence base, FDI can be given in single high-dose infusions and is supported by recent outcome trials (IRONMAN), while IS requires multiple small infusions but remains widely available in many regions due to lower cost. Direct head-to-head RCTs are lacking, and formulary access varies globally; therefore, a comparative framework is clinically important to inform practice and guideline recommendations. Our analysis provides an updated comparative framework, integrating results from recent large-scale outcome trials and prior meta-analyses.

### 4.1. Principal findings and comparison with previous studies

We found that intravenous iron supplementation significantly reduced the risk of heart failure hospitalization, with the most robust evidence for FCM (high certainty) and supportive but less conclusive evidence for FDI and IS. These results align with prior meta-analyses [2,7,8,9,10,11] and major RCTs such as AFFIRM-AHF [4], which demonstrated a 21% relative reduction in recurrent HF hospitalizations with FCM, and IRONMAN [5], where FDI showed a consistent, though not statistically significant, trend toward fewer hospitalizations. The recent HEART-FID trial [6], the largest to date, did not meet its primary endpoint, but its findings of reduced HF hospitalizations are directionally consistent with our pooled results. FAIR-HF2 [23] further supports the role of FCM in improving functional outcomes without clear mortality effects.

In contrast, no mortality benefit was observed for any formulation. Neither all-cause nor cardiovascular mortality was significantly reduced, consistent with previous meta-analyses [7,9–11] and individual trial results [4–6,14,17]. The lack of mortality reduction may reflect limited statistical power, short follow-up, or a true absence of survival benefit despite improvements in morbidity.

Regarding functional capacity, all three formulations improved 6MWD, with the largest average gains seen with IS and FDI, though accompanied by high heterogeneity (I^2^ >90%). These improvements are consistent with earlier efficacy trials such as FAIR-HF [14], CONFIRM-HF [17], and EFFECT-HF [18]. However, as highlighted in recent work on minimal clinically important differences [27], the average improvement of ∼20–30 meters with FCM may be below the threshold for clear clinical relevance, underscoring the importance of patient-level contextualization.

Follow-up periods varied widely across studies, ranging from 12 weeks (e.g., EFFECT-HF, AFFIRM-AHF) to ∼1 year (HEART-FID) and over 2 years in IRONMAN (median 2.7 years) [5,6,18]. This variability in duration likely contributes to observed heterogeneity for continuous outcomes and limits conclusions about long-term mortality effects.

As expected, iron indices improved substantially with all formulations. FDI was associated with the greatest ferritin increases, while FCM demonstrated the most consistent improvements in TSAT. These findings mirror prior small RCTs [12,13,15,16,19,20] and confirm the pharmacologic capacity of IV iron to correct iron deficiency more effectively than oral supplementation. Across trials, FCM was generally administered in 500–1000 mg doses at baseline with repeat dosing after 1–6 weeks to reach cumulative doses of 1500–2000 mg [14,17,18]. FDI was typically given as a single infusion of 1000–1500 mg, with repeat dosing during long-term follow-up (e.g., IRONMAN) [5,20]. IS required multiple 200 mg infusions, often 5–10 sessions, to achieve repletion [12,13], making it less convenient in clinical practice.

Patient-reported outcomes such as KCCQ scores and safety endpoints (adverse events, serious adverse events) were not included in this analysis due to insufficient and inconsistent reporting across studies. While individual trials such as CONFIRM-HF did evaluate KCCQ [17], the lack of standardized outcome definitions prevented reliable synthesis. Future research should better capture quality of life and safety endpoints to complement clinical outcomes.

### 4.2. Policy and clinical implications

Current guidelines emphasize the role of IV iron in HFrEF. The 2021 ESC guidelines recommend FCM for symptomatic patients with LVEF <45% and iron deficiency (Class IIa, Level A) [3]. The 2022 ACC/AHA/HFSA guidelines and the 2023 HFSA scientific statement [1] similarly endorse IV iron therapy, particularly FCM, to improve quality of life and reduce HF hospitalizations. Our findings support these recommendations, as FCM remains the formulation with the largest and most robust evidence base. Evidence for FDI is promising, particularly after IRONMAN [5], but additional trials are needed to consolidate its role. Data for IS remain limited to smaller, earlier studies [12,13], and although efficacy signals exist, logistical challenges (multiple smaller infusions) may reduce its clinical attractiveness compared with newer formulations.

Clinicians should prioritize IV iron therapy in iron-deficient HFrEF patients for morbidity reduction, while acknowledging that mortality benefits remain uncertain. Tailored approaches may be considered in acute versus chronic HF settings and in patients with concomitant chronic kidney disease, but current evidence is strongest in ambulatory HFrEF populations with reduced LVEF ≤45%.

## 5. Study limitations

This study has several limitations. First, heterogeneity was substantial for continuous outcomes such as 6MWD, ferritin, and TSAT, reflecting variation in baseline populations, trial designs, and follow-up durations. Second, although funnel plot inspection suggested possible small-study effects, the limited number of available trials per outcome reduced power for formal bias testing. Third, no head-to-head RCTs directly comparing FCM, FDI, and IS exist; thus, all formulation-level comparisons rely on indirect evidence. Fourth, several outcomes—particularly ferritin and TSAT—represent surrogate markers of iron repletion rather than hard clinical endpoints, and their clinical significance must be interpreted with caution. Finally, the majority of included patients had HFrEF with EF ≤40–45%, limiting generalizability to HFpEF populations.

## 6. Conclusions

In patients with HFrEF and iron deficiency, intravenous iron supplementation—particularly with ferric carboxymaltose—significantly reduces heart failure hospitalization and improves functional capacity, while mortality benefits remain uncertain. All formulations effectively replenish iron stores, with FDI showing the largest ferritin gains and FCM the most consistent improvements in TSAT. These findings strengthen the role of IV iron as an adjunctive therapy in HFrEF, as reflected in current guideline recommendations. However, further high-quality RCTs of FDI and IS, longer-term trials powered for mortality, and studies in HFpEF and real-world settings are warranted to refine treatment strategies and expand generalizability.

## Data Availability

All data produced are available online at PubMed.

## Authors’ contributions

All authors contributed equally to the manuscript and read and approved the final version of the manuscript.

