## Supplemental figures and Appendix for "Comparative Effectiveness of Intravenous Ferric Carboxymaltose, Ferric Derisomaltose, and Iron Sucrose in Heart Failure With Reduced Ejection Fraction: A Network Meta-Analysis of 15 Randomized Trials"

#### Supplementary Figure 1. Risk of bias assessments (traffic-light and bar plots).

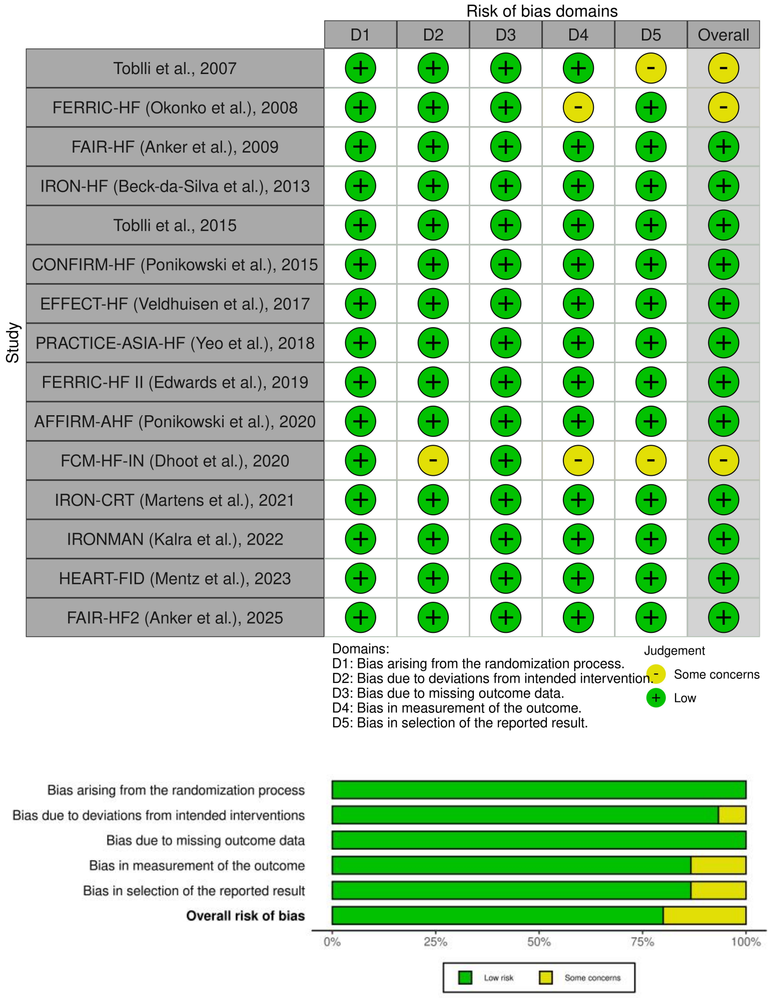


#### Supplementary Figure 2. Funnel plots for publication bias across outcomes.

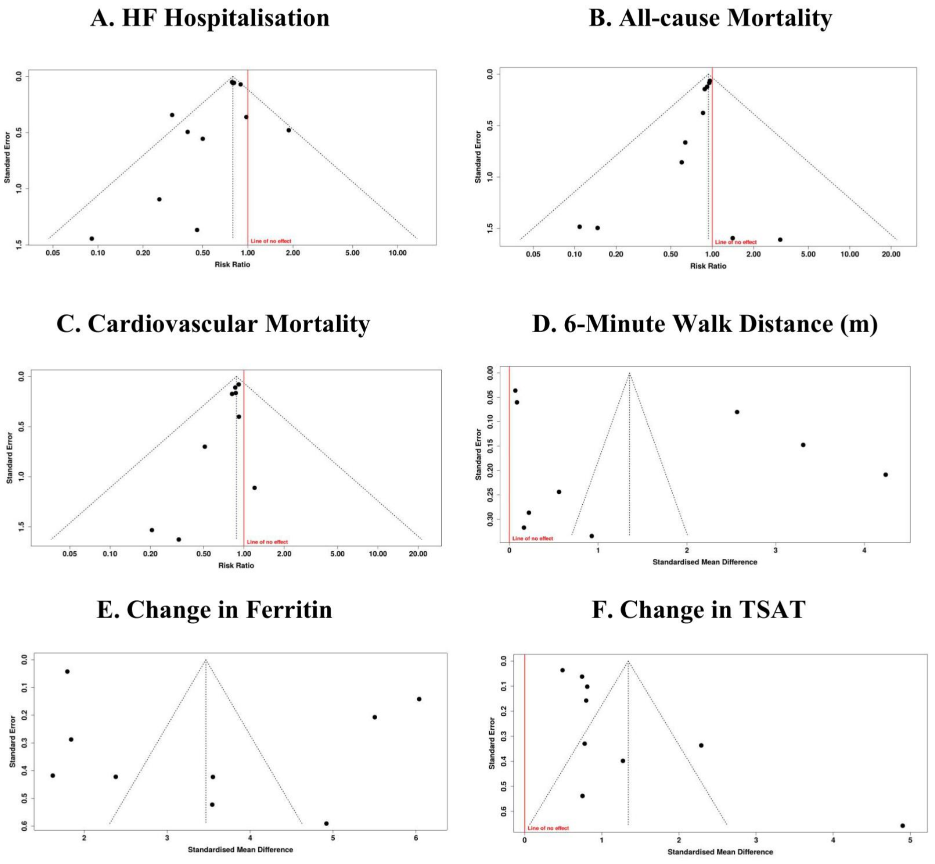

#### Supplementary Figure 3. Separate forest plots of individual trial results by outcome.

##### B. All-cause Mortality

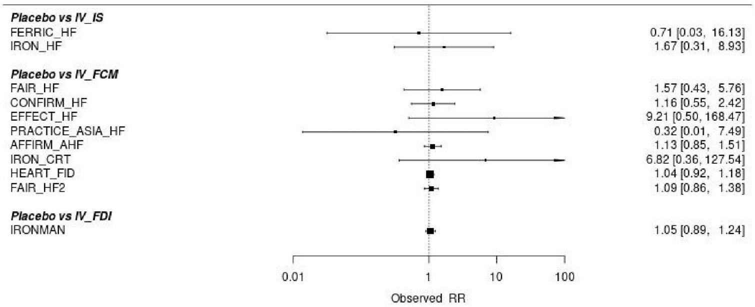


##### C. Cardiovascular Mortality

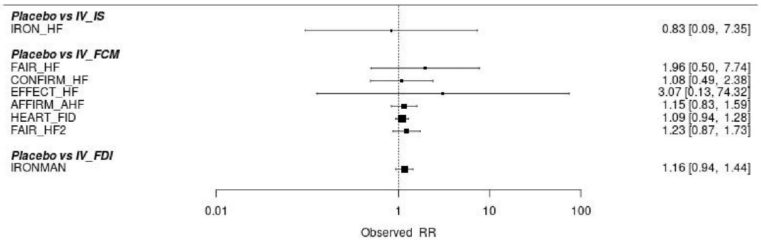


##### 6-Minute Walk Distance (m)

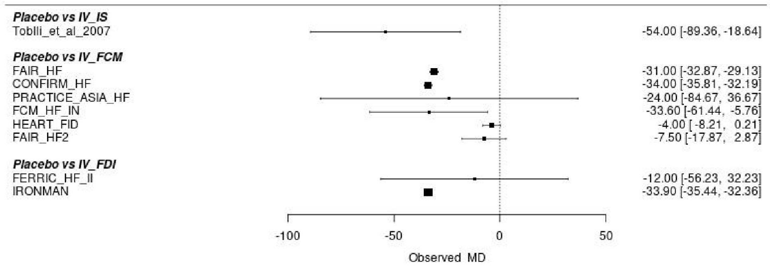


##### E. Change in Ferritin

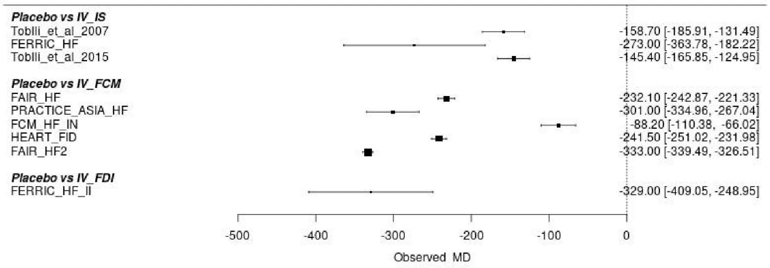


##### F. Change in TSAT

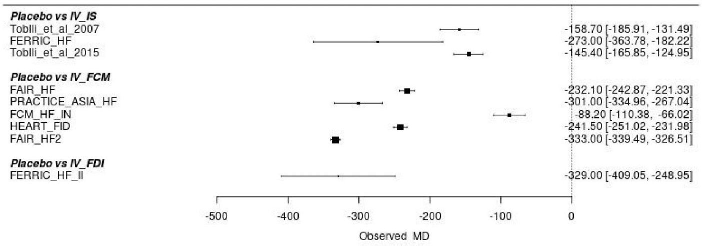


#### Supplementary Figure 4. Meta-regression plots for continuous outcomes.

##### A. HF Hospitalisation

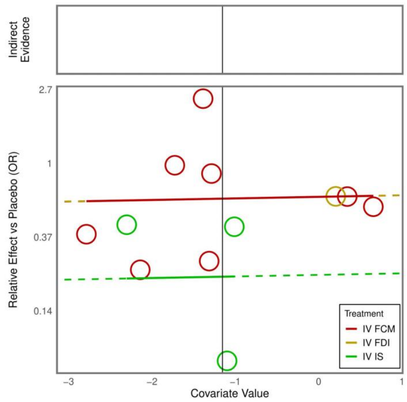


##### B. All-cause Mortality

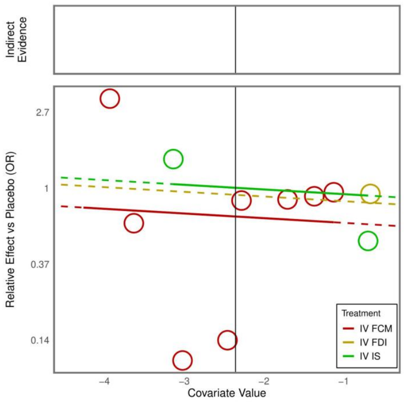


##### C. Cardiovascular Mortality

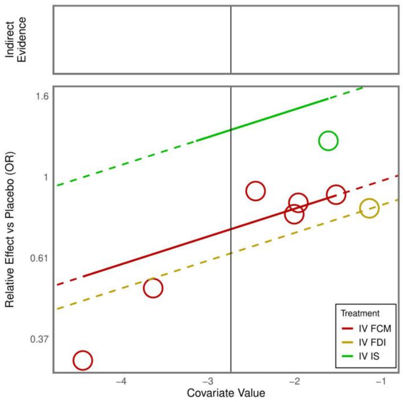


##### D. 6-Minute Walk Distance (m)

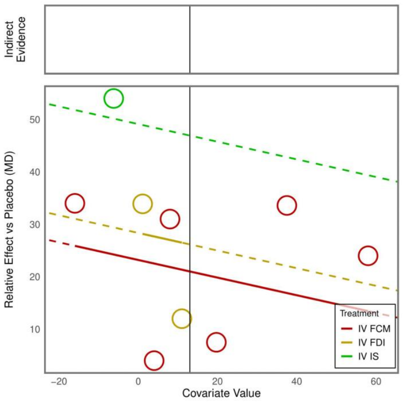


##### E. Change in Ferritin

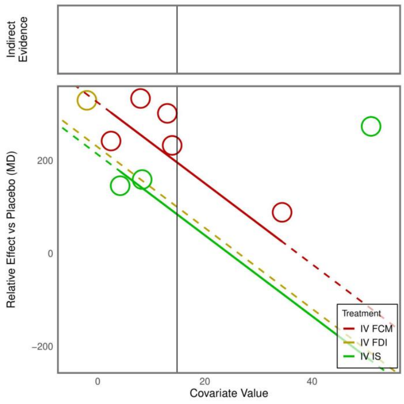


##### F. Change in TSAT

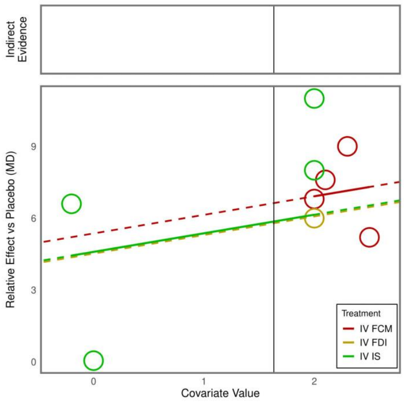

### Appendix 1

PubMed/MEDLINE (via MeSH + free text):

("Heart Failure"[MeSH] OR "cardiac failure" OR "systolic heart failure") AND ("Iron Deficiency"[MeSH] OR "iron deficiency" OR "iron-deficient") AND ("Ferric Carboxymaltose" OR "Carboxymaltose, Ferric"[MeSH] OR "Ferric Derisomaltose" OR "Iron Isomaltoside" OR "Iron Sucrose" OR "Sucrose, Iron"[MeSH] OR "intravenous iron" OR "IV iron") AND (randomized controlled trial[pt] OR randomized[tiab] OR placebo[tiab])

CENTRAL (Cochrane Library):

#1 "Heart Failure" OR "cardiac failure" #2 "Iron Deficiency" OR "iron-deficient" #3 "Ferric Carboxymaltose" OR "Ferric Derisomaltose" OR "Iron Isomaltoside" OR "Iron Sucrose" OR "intravenous iron" #4 #1 AND #2 AND #3 in Trials

Web of Science (Topic search):

TS = ("heart failure" OR "cardiac failure") AND TS = ("iron deficiency" OR "iron-deficient") AND TS = ("ferric carboxymaltose" OR "ferric derisomaltose" OR "iron isomaltoside" OR "iron sucrose" OR "intravenous iron") AND TS = ("randomized" OR "placebo")
